# RESCUE: An end-to-end multi-agent LLM system for proactive rare-disease patient screening in the EHR

**DOI:** 10.64898/2026.06.24.26356357

**Authors:** Cong Liu, Alexa Geltzeiler, Adam Afyouni, Mengshu Nie, Kamalakkannan Ravi, Courtney French, Monica Wojcik, Wendy K. Chung

## Abstract

Rare diseases affect a significant portion of the global population, yet patients often endure a lengthy diagnostic odyssey and miss timely diagnosis with exome or genome sequencing (ES/GS). Existing informatics tools often rely on pre-identified patients or institution-specific rule sets that are laborious to implement locally and rarely address clinical utility and operational feasibility. We introduce RESCUE, an end-to-end, multi-agent LLM-powered workflow that automates the local implementation of both rule-based and machine learning screening algorithms across the entire electronic health record (EHR). Four specialized agents operate in sequence: the Ontology Agent classifies clinical data into a four-tier genetic-evidence taxonomy; the Modeling Agent builds a positive-unlabeled (PU) XGBoost classifier; the Screening Agent applies the model across the EHR and flags patients with a rule-based algorithm; and the Review Agent samples clinical notes to assess medical necessity and operational feasibility. In a retrospective evaluation at a pediatric hospital, the model showed strong discrimination on a holdout set (n=12,591; AUC 0.808). Of 175,842 patients meeting screening criteria, those flagged by RESCUE were 7.4-fold more likely to receive subsequent genomic assessments than controls. Blinded chart review of this group confirmed previously missed, medically appropriate candidates for ES/GS with 80% precision, accounting for prior testing. By decoupling expert roles into modular agents, RESCUE offers a flexible, scalable framework, with machine-learning and rule-based approaches complementing rather than substituting for each other. It also provides a reproducible, agentic pathway for direct local implementation to reduce diagnostic delays on an institutional scale.

## 1. Introduction

Rare diseases collectively affect 3.5∼6% of the population, spanning more than 7,000 distinct disorders, and most are genetic in origin^1^. For suspected Mendelian diseases, exome and genome sequencing (ES/GS) yields molecular diagnosis in 25–50% of suspected cases^2–4^ and is always recommended as a first or second-tier test for congenital anomalies, developmental delay, intellectual disability, and many other indications^5,6^. Even so, the average time between first symptoms and a diagnosis is 5∼7 years^7^, and most who could benefit are never referred to a clinical geneticist ^8^. Together, these observations point to a recognition gap rather than a testing gap, where the limiting step is identifying which patients should be offered genomic sequencing.

Most existing tools assume the patient has already been identified. Ontology-based pipelines^9,10^ and gene-prioritization tools such as Phen2Gene^11^, Phen2Disease^12^, PheNet^13^, Phenomizer^14^, Phenolyzer^15^, Exomiser^16^, and AMELIE^17^ all start from a phenotype list a clinician has already assembled for a patient who is undergoing analysis. Recently, ThinkRare^18^ was developed to scan the EHR with a hand-crafted rule set; because its rules are tied to one institution’s coding taxonomy, adopting it at other sites requires local reconstruction. Furthermore, none of these tools address the essential operational question: would this patient benefit from genomic testing, has the diagnostic work-up already been completed, and has the patient been assessed by a geneticist?

Large language models (LLMs) provide the potential to change this landscape. LLM can now read clinical notes at close to expert level^19^, and extract phenotype concepts out of free text^20,21^. Additionally, they can act as *agents:* calling tools like database queries, code execution, and model training^22,23^, implementing previously published algorithms, and reviewing charts to carry out multi-step tasks. This agentic capability is a good fit for EHR based rare-disease screening, where the workflow draws on many pre-validated tools and must interact with several separate digital systems.

In this study we introduce **RESCUE (***Rare Disease Detection and Escalation Support via a Learning Health System*), an EHR-screening *agentic workflow* for proactive identification of patients who are likely to have rare diseases and would benefit from clinical genomic sequencing. RESCUE attempts to automate the local implementation of such algorithms rather than replace any rule-based or machine-learning models. It is a multi-agent system consisting of multiple LLM-powered agents, each with a skill inventory including free-text description and pre-validated python script. To our knowledge, RESCUE is the first agentic workflow to run end to end at EHR scale: building the cohort, training a machine-learning model, implementing a rule-based algorithm, screening patients with both, and reviewing charts under operational constraints.

## 2. Methods Study setting

RESCUE was developed and evaluated using the i2b2 enterprise data warehouse^24^ (hosted on Snowflake) of a large tertiary pediatric hospital. Each agent is powered by a customized OpenAI Codex CLI, with all LLM calls directed to a HIPAA-compliant intramural Azure OpenAI endpoint. To ensure patient privacy, tracing is disabled, and the Codex’s web browsing capabilities are turned off to prevent accidentally sending protected health information (PHI) to external sources. By employing these measures, no PHI leaves the institutional firewall. This study has been approved by the Boston Children’s Hospital institutional review board. The data used for this study will NOT be published due to patient-sensitive information.

### Agent architecture

An overview of the multi-agent RESCUE design and evaluation is shown in Figure 1. RESCUE is implemented as four agents (with one additional agent reserved for evaluation in this study; see *Evaluation*). Each agent has a defined goal, a fixed set of skills, and a single canonical output. Users can control the configuration file (or use another agent called “Vibe Coding” to modify it) to provide details of the parameters, such as screening date and index window. The agents were chained by an orchestra by running in order, with each agent’s output feeding the next. Every long running call is checkpointed row by row with results cached locally after each patient is processed, so an interruption resumes from where it stopped without repeating completed API calls. We provide a brief description of each agent below. The detailed agent settings can be found at https://github.com/stormliucong/RESCUE-Agent.

**Figure 1.**
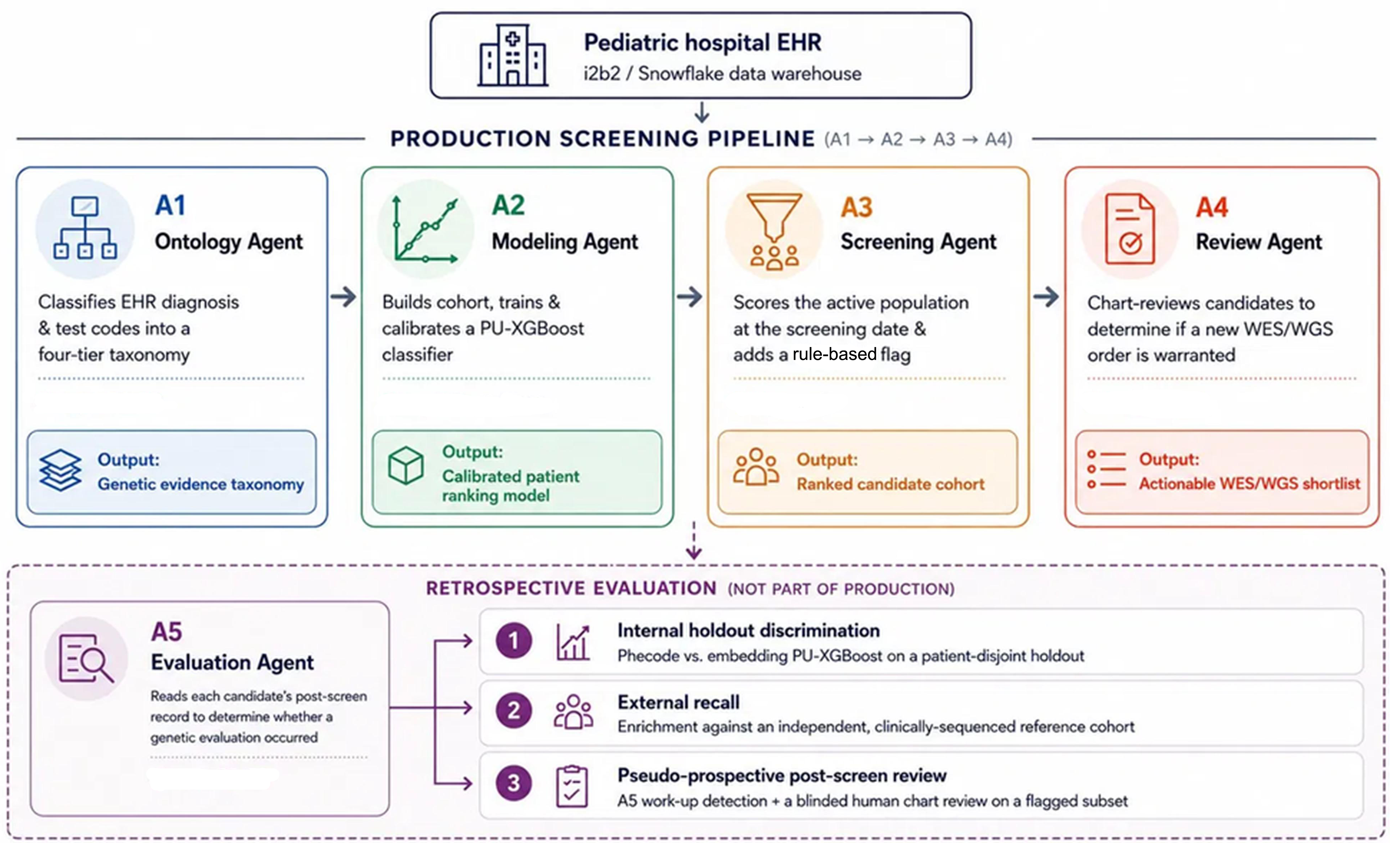
Overview of the multi-agent RESCUE design and evaluation studies. The Ontology Agent (A1) classifies EHR diagnosis and test codes into a four-tier genetic-evidence taxonomy; the Modeling Agent (A2) builds a case/control cohort and trains a calibrated PU-XGBoost classifier; the Screening Agent (A3) scores the active patient population at the screening date and adds a flag from a reconstructed rule-based algorithm; and the Review Agent (A4) chart-reviews candidates to produce the final eligible list for a new ES/GS order. A1–A4 run in production; the Evaluation Agent (A5) is used only retrospectively, to check whether flagged patients later underwent genetic evaluation.

### Agent A1: Ontology Agent

Goal: turn the raw EHR concept dictionary into a four-tier genetic-evidence taxonomy. Skills:

- A Snowflake query to query i2b2 over OBSERVATION_FACT (i.e., the primary table describing all the clinical details of patients) and CONCEPT_DIMENSION (i.e., a dictionary mapping each clinical code (i.e., ICD code, test code) to the specific descriptions) to enumerate every ICD code and every keyword-flagged test concept.
- Instructions and examples to classify each ICD or internal test code into a predefined semantic category based on code description and their metadata.

Outputs:

- **Tier 1A** — ICD codes classified as genetic indications (risk-enrichment).
- **Tier 1B** — non-diagnostic / screening / pharmacogenomics-related genetic tests (control-purification).
- **Tier 2A** — test codes classified as diagnostic genetic tests (case-defining).
- **Tier 2B** — ICD codes classified as established genetic diagnoses (case-defining).

### Agent A2: Modeling Agent

Goal: assemble a positive-unlabeled cohort with matched controls based on the class labels produced by A1; construct patient feature matrices from pre-index diagnostic history and train a calibrated PU-XGBoost screening model.

Skills:

- A Snowflake query to extract 300-character windows around ES/GS keyword mentions in clinical notes and classify whether each mention reflects a rare-disease diagnostic context.
- A Snowflake query to assemble a positive-unlabeled (PU) cohort (cases = Tier 2A/2B or ES/GS-in-context patients; unlabeled pool purified of all Tier 1/2 exposure), with one-to-many matching by age band and per-year visit count.
- Instructions to construct two parallel feature representations from pre-index ICD-10 codes, excluding case-defining Tier 2A/2B codes to prevent leakage: (a) a binary Phecode matrix with visit aggregates and age at index, and (b) dense 3,072-d vectors via text-embedding-3-large applied to a structured serialization of each patient’s diagnostic history.
- A validated Python template to train a PU-XGBoost classifier on both feature sets with a patient-level 80/20 stratified holdout and isotonic calibration.

Outputs:

- Cohort for PU learning include training/validation and holdout sets
- PU trained models and metadata (performance) including a Phecode model and an embedding model.

### Agent A3: Screening Agent

Goal: apply the trained model (by default Phecode-based) at a fixed screening date and produce a ranked list of candidate patients, alongside a flag from a rule-based algorithm (described below).

Skills:

- A Snowflake query to enumerate the institutional patient base (∼5 million) at the screen date and apply the same inclusion filter used during training (no prior Tier 2A (diagnostic genetic test) or Tier 2B (confirmed diagnosis) code; ≥2 visits in each of the 2 pre-screen years).
- A validated Python template to re-extract pre-screen Phecode features under the metadata of trained models (missing columns zero-filled), calls the calibrated PU model, and sorts by calibrated probability.
- Instructions and original paper to implement a rule-based algorithm reconstructed from the ThinkRare rules^18^ and adapted to our EHR schema. Note that our implementation is not equivalent to the original ThinkRare implementation since some exclusion rules are not included in our approach; we therefore refer to it as the rule-based algorithm throughout.

Outputs:

The output is one row per screened patient with raw probability from the model, calibrated probability that estimates the likelihood that the patient whose diagnostic history resembles that of the positive class patients and a Boolean label for the rule-based flag (rule-flagged vs. rule-unflagged).

### Agent A4: Review Agent

Goal: Determine whether ES/GS would be a necessary clinical action for each candidate from A3’s ranked list. In production, the Review Agent runs only on top-ranked candidates; for the retrospective evaluation, parallel control arms were also processed (see Evaluation).

Skills:

- A Snowflake query that returns metadata-only summaries of each candidate’s pre-screen notes.
- A weighted-sampling policy that selects up to 10 notes per candidate, with a fixed high-priority weight (=99) for notes with text that contains any of *genetic, exome, genome, sequencing, chromosome, chromosomal, ES, GS, microarray*, and an exponential-decay × note-type-boost weight for all other notes.
- A technique to reduce LLM API calls per patient by pulling only the sampled note blobs and concatenating them.
- Instructions to classify note blobs to determine prior genetic testing status, and exclude patients with a prior positive genetic test, prior ES/GS testing, or multiple unfulfilled genetic test orders (for any reason).
- Instructions to assess medical necessity against ACMG clinical criteria and filter out patients who do not meet them (configurable for evaluation).

Outputs:

- A final list of patients for whom ES/GS was found to be medically necessary, and operationally feasible.

### Agent A5: Evaluation Agent (not for production)

Goal: For each candidate produced in A4, read what happened in their chart after the screen date and determine prospectively whether they actually went on to have a genetic evaluation. This is only used to study evaluation purposes.

Skills:

- A Snowflake query example to flags candidates with enough follow-up to judge. “Enough” means at least 2 post-screen visits, with the last visit at least 183 days (about 6 months) after the screen date.
- A validated Python template to look for a post-screen genetic workup through two channels: a structured channel (any Tier 2A or 2B observation recorded after the screen date) and an unstructured channel (the LLM reading the patient’s post-screen notes, see *system prompt P5* in Supplementary Appendix).
- A validated Python template to sample notes by reusing A4’s weighted sampling but flips the time weight so recent notes are favored (1 - exp(-days/365)), gives genetic-keyword notes a high fixed weight, and caps it at 10 notes per patient.
- Instructions to perform chart review on sampled notes per patient and to answer the question: *did the patient get a genetic workup after the screen?* (*Supplementary Appendix prompt P8*)

Outputs:

- A list of candidates who had a post-screen genetic workup. In the case arm (the patients the screen flagged) these are true positives; in the control arm they are false negatives, since the screen missed them.
- A list of candidates who had enough follow-up but no workup found in either channel. These are sent for manual review in the evaluation study.

### Evaluation

We performed three evaluation experiments to benchmark RESCUE:

1. Internal holdout discrimination. Agent scored the Phecode and embedding PU-XGBoost classifiers on a patient-disjoint 20% holdout. Metrics reported include AUC (area under the ROC curve), AP (average precision), calibration, and stratified odds ratios (ORs) across cases, risk-enriched patients, and purified controls.
2. To evaluate whether RESCUE recognizes rare-disease patients in the absence of explicit coding, we tested it against the Children’s Rare Disease Cohort^25^ (CRDC), an independently curated registry of patients confirmed to have rare diseases through clinical genomic sequencing. Since the CRDC was outside of the coded EHR data used for RESCUE training, it serves as an external ground truth for recall. We pass this cohort through the same patient-inclusion filter, scored the model on every patient, and measured enrichment of CRDC members across configurable cutoffs. Recall and odds ratios were therefore computed within this RESCUE-eligible population rather than the full CRDC cohort.
3. Pseudo-prospective (post 2021-12-31) evaluation, comprising a review agent (A5) that adjudicates whether a candidate underwent genetic evaluation after the screen date; and a blinded human chart-review on a subset of cases and controls flagged by A5 as having adequate follow-up but no detected genetic workup after the index date.

## 3. Results

The Ontology Agent classified 14,000 high-prevalence ICD-10 codes and 1,200 keyword-flagged test codes into the four-tier framework. The number of included codes in each tier can be found in *Supplementary Appendix Table S1*. The training and validation cohort thus came to 6,153 unique cases and 56,799 unique controls using the following inclusion criteria: index date = first qualifying event shifted backward 90 days; index window 2018-01-01 to 2021-12-31; activation requires ≥2 visits in each of 2 lookback and 2 follow-up years. The Modeling Agent trained the Phecode and embedding PU-XGBoost classifiers on the same 50,361 training patients and evaluated them on a 12,591-patient holdout (1,230 cases, 11,361 controls). As shown in ***Table 1***, both Phecode model (AUC = 0.794, AP = 0.400) and Embedding model (AUC = 0.808, AP = 0.417) achieved comparable performance (*supplementary Figure S1*) and isotonic calibration left both curves close to the diagonal (*supplementary Figure S2*). We also checked whether the model picked up patients carrying genetic indication codes (Tier 1A, the risk enrichment tier). At every non-zero cutoff, flagged patients were enriched for these codes (*supplementary Tables S2*; *Figure 2*). At a calibrated cutoff of 0.20, for example, the Phecode model flagged 33.1% of patients who had a genetic indication code versus 8.3% of those who did not (OR 10.1).

**Figure 2.**
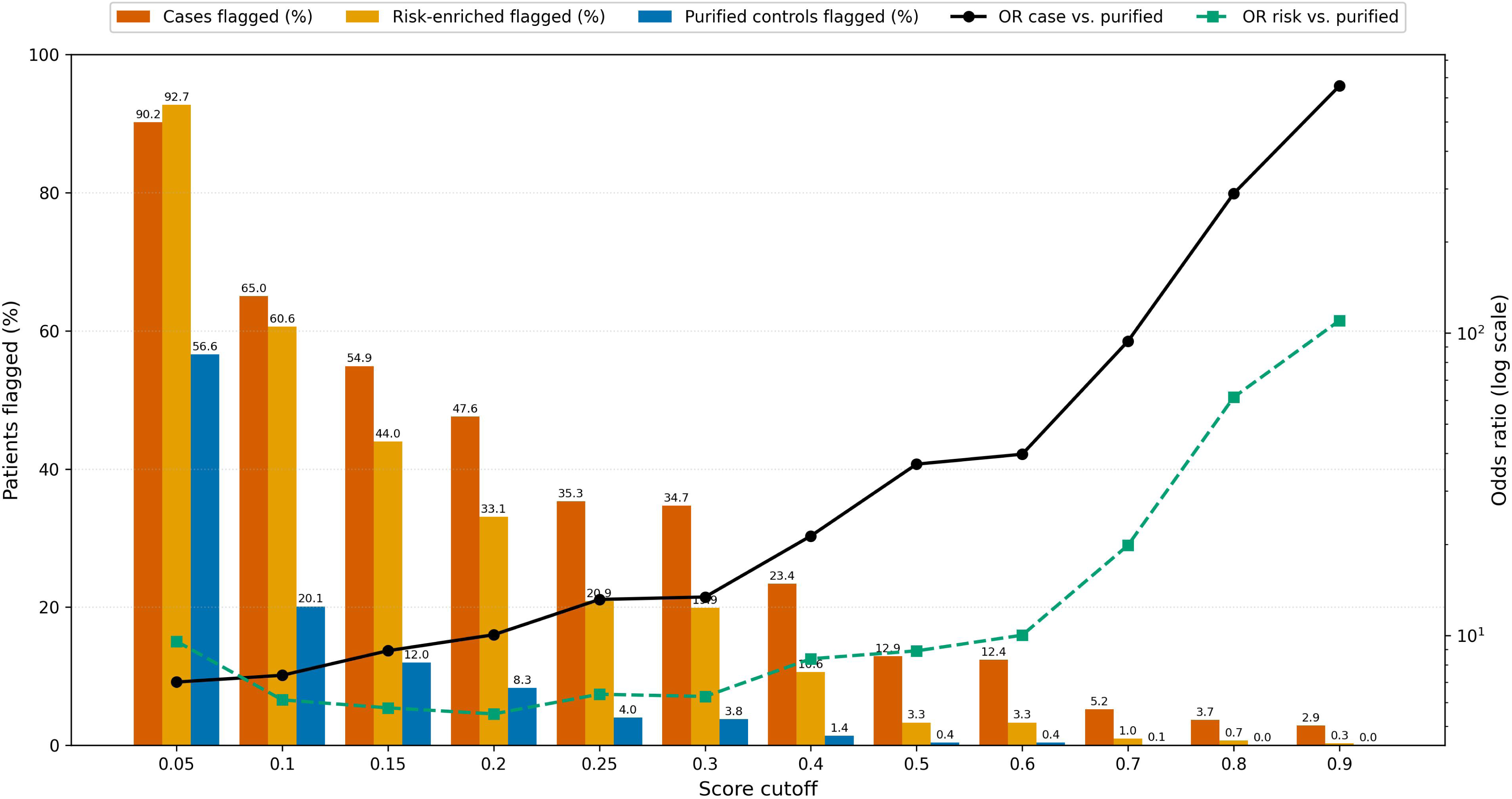
Patients flagged in different groups across model score cutoffs. Grouped bars show the percentage of patients flagged (i.e., with a predicted score at or above the cutoff) within each group: cases (vermillion), risk-enriched patients (orange), and purified controls (blue). Data labels above each bar indicate the exact percentage flagged. Overlaid lines show the odds ratio (OR) of being flagged for cases versus purified controls (black, solid) and for risk-enriched patients versus purified controls (green, dashed).

**Table 1.** Comparison of Phecode and embedding-based PU-classifier on Holdout validation dataset.

| **Model** | **N features** | **Per record (Row) AUC** | **Per record (Row) AP** | **Patient AUC** | **Patient AP** |
|---|---|---|---|---|---|
| Phecode | 1,616 | 0.794 | 0.400 | 0.796 | 0.389 |
| Embedding | 3,072 | **0.808** | **0.417** | **0.811** | **0.404** |

AP: average precision score. AP summarizes a precision-recall curve as the weighted mean of precisions achieved at each threshold, with the increase in recall from the previous threshold used as the weight: AUC: Area Under the Receiver Operating Characteristic Curve (ROC AUC) from prediction scores. Per record: each unique patient in unlabeled group can have more than records (i.e. different index date)

The Screening Agent then applied the trained models across the full institutional EHR at a screen date of 2021-12-31. Among 494,577 active patients, 97,021 known cases (Tier 2A/B prior to screen; see *Table S1*) and 268,438 patients with insufficient pre-screen visits were excluded, leaving 175,842 patients meeting the inclusion criteria. The mean calibrated positive prediction probability is 0.077 among those included patients, and the rule-based algorithm flagged 2,678 (1.5%) of these patients. *Supplementary Table S3* shows the calibrated-probability cutoff breakdown and the overlap between the PU-trained model and the rule-based algorithm. Even at probability ≥0.05 only 2.8% of model-positives were rule-flagged, indicating that the two methods carry largely complementary rather than redundant signals.

We also benchmarked the Screening Agent against the Children’s Rare Disease Cohort (CRDC; n = 8,074 sequenced patients), passing it through the same inclusion filter the agent uses. That left 2,049 CRDC patients eligible at 2021-12-31; the recall and OR estimates below are computed within this RESCUE-eligible population rather than the full CRDC cohort. At probability ≥ 0.20 with rule-flagged patients (i.e., the intersection of both approaches), recall was 3.5% and the OR against eligible non-CRDC patients was 4.8. (*Table S4; Figure 3*).

**Figure 3.**
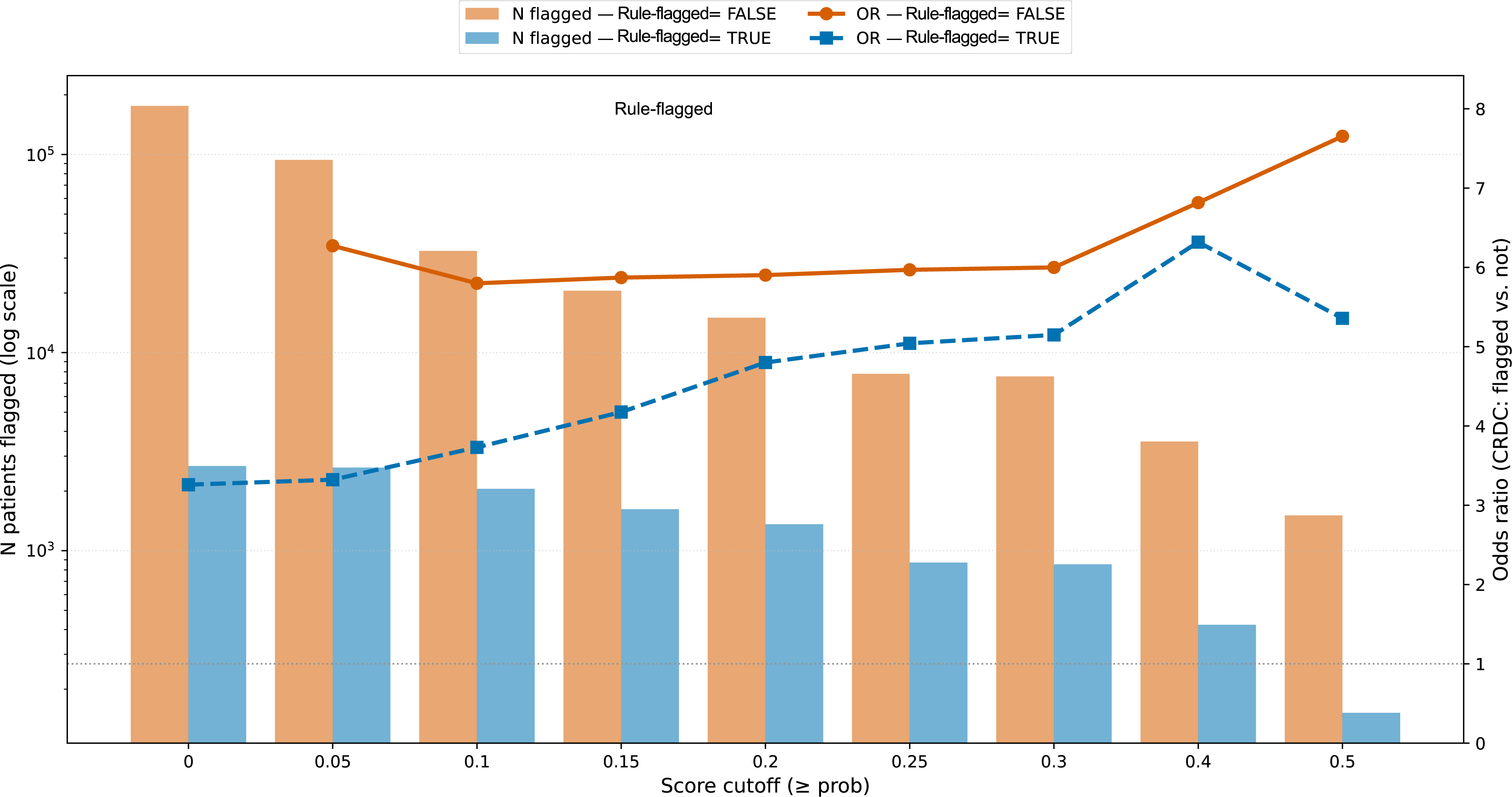
Screening performance based on CRDC cohort. Patients were stratified by whether they were independently flagged by the rule-based algorithm (rule-flagged; n = 2,678; blue) or not (rule-unflagged; n = 175,842; vermillion), and the RESCUE model was evaluated separately within each stratum across calibrated prediction probability cutoffs. Bars (left axis, logarithmic scale) show the number of patients flagged at or above each cutoff. Lines (right axis) show the odds ratio of being a confirmed rare disease case (CRDC, patients who have received ES/GS) among flagged versus non-flagged patients at each cutoff; the dotted horizontal line marks an odds ratio of 1 (no enrichment). Recall and odds ratios are calculated within the 2,049 RESCUE-eligible CRDC patients rather than the full CRDC cohort.

In production the Review Agent runs only on the top-ranked patients, although this can be customized. For evaluation, we tried four different configurations by pointing them at four comparison arms defined by the PU-trained model probability (M) and the rule-based flag (R):

- **M+R+**: calibrated probability >0.5 AND rule-flagged (n = 152).
- **M-R+**: calibrated probability ∈ (0.05, 0.10) AND rule-flagged (n = 577)
- **M+R-**: calibrated probability >0.5 AND rule-unflagged, capped at 1,000 random sampled patients.
- **M-R-**: calibrated probability ∈ (0.05, 0.10) AND rule-unflagged, capped at 1,000 random sampled patients.

The Review Agent flagged pre-screen prior-testing evidence in 31/152 (20.4%) of M+R+ candidates, 49/577 (8.5%) of M-R+ candidates, 204/1,000 (20.4%) of M+R-candidates, and 32/1,000 (3.2%) of M-R-candidates. After A4 removed candidates with documented prior testing, 121 M+R+, 796 control M+R-, 528 M-R+, and 968 M-R-candidates remained in the operationally actionable lists. The Evaluation Agent then ran over the follow-up-eligible patients in each arm to see who had a post-index genetic workup (*Table S5*; *Figure 4*). Patients in the M+R+ arm (i.e., selected by both the PU-trained model and the rule-based algorithm) were 7.4-fold more likely than those in the M-R-arm (i.e., low model probability, rule-unflagged) to have documented genetic-evaluation activity (26.5% vs. 3.6%).

**Figure 4.**
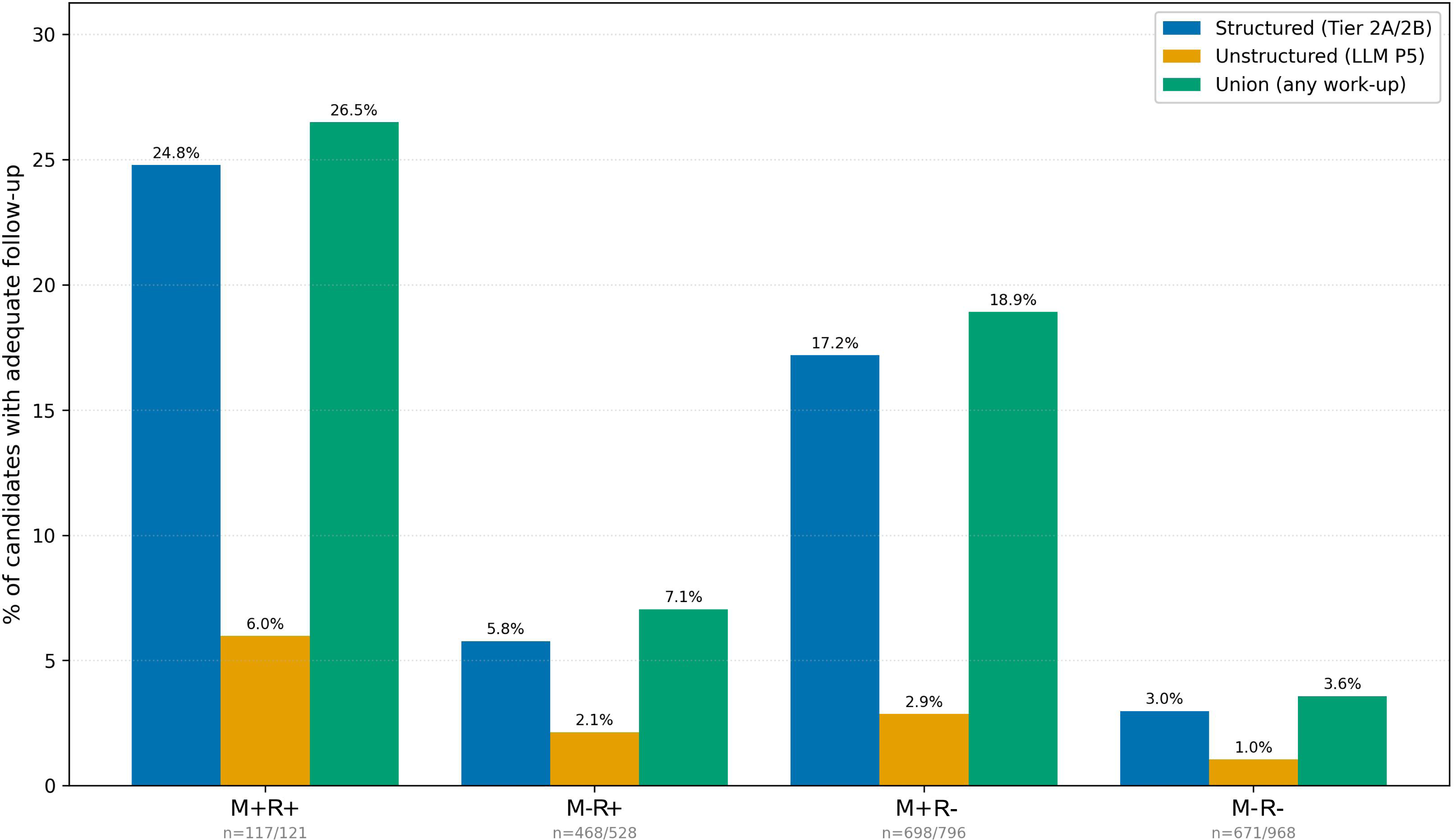
Post-screening evaluation across arms. Candidates were partitioned into four mutually exclusive arms by whether they were flagged by the PU-trained model (M+/M−) and whether they were flagged by the rule-based algorithm (R+/R−). For each arm, bars show the percentage of candidates with adequate follow-up who had evidence of a diagnostic genetic work-up ordered, by evidence source: structured evidence (Tier 2A/2B diagnostic codes; blue), unstructured evidence (LLM-identified work-up in clinical notes; orange), and the union of any work-up from either source (green). Percentages are expressed relative to the number of candidates with adequate follow-up in each arm.

Finally, an independent board-certified genetic counselor did a blinded chart review of a 15-patient subsample for the M+R+ and M-R-arms, all flagged by the Evaluation Agent as having no detected post-index workup. In the M-R-arm, the counselor agreed that 14/15 (93.3%) had no genetic relevance or medical necessity to order ES/GS. In the M+R+ arm, 13/15 (86.7%) met medical necessity criteria for testing; one of those had a prior order the Review Agent missed.

That left 12/15 (80.0%) M+R+ patients judged both medically necessary and operationally actionable, that is, worth contacting the patient or provider for genomic testing.

## 4. Discussion

Screening electronic health records (EHRs) to identify patients who are appropriate candidates for exome sequencing (ES) or genome sequencing (GS) traditionally involves multiple areas of expertise. This includes EHR coding terminology experts, professionals who can train and validate data-driven models, and genetic specialists who review charts to ensure accuracy and operational feasibility. RESCUE applies this approach by assigning the roles of coding terminology expert, machine learning engineer, and genetic counselor to separate agents, each responsible for one specialized part of the pipeline. By adopting a multi-agent design^26^, each agent is given a narrow and defined scope, making each task easier to specify and the output easier to constrain. The retrospective analysis facilitated by Evaluation Agent (A5) indicates that individuals flagged by both PU-trained model and the rule-based algorithm without prior genetic evaluation had a significantly elevated likelihood of ultimately receiving genetic evaluation and testing.

As an agentic workflow, RESCUE provides a straightforward way to implement existing rule-based algorithm directly within a local institution. Our data suggest that, although the ThinkRare¹ rules were developed around the coding taxonomy of a different institution, a rule-based algorithm reconstructed from its published description and adapted to our local EHR still performed well, indicating that the underlying approach may be transferable to other sites. We overcome the challenge of the absence of labeled datasets for supervised learning in rare diseases^27^ by employing a positive-unlabeled (PU) learning approach. The Ontology Agent eliminates the need for manual review of institution-specific codes, producing proxy labels for training. Although the actual code may not be easily transferable due to variations in data schemas and internal codes across institutions, the ontology agent and corresponding prompts are expected to be readily adaptable to other clinical settings with minimal changes. Similar to other LLM-powered chart review^28^, The Review Agent adds an operational-feasibility check by sampling the most informative notes to determine if genomic testing has already been ordered and if the patient is operationally reachable. This allows clinically deployed systems to weigh logistical concerns such as ability to contact the patient and family and prior work-up alongside clinical justification. Each clinic can further revise the prompt to encode its own operational rules, enhancing the precision of the flagged list.

Our evaluation demonstrates that the Phecode-based^29^ approach and the embedding-based^30^ approach perform similarly, suggesting that a large transformer-based encoding approach can capture much of the information in human-curated ontology systems. The more operational decision is where to set the screening cutoff, which should be tuned to match each institution’s recontact capacity. A lower threshold maximizes recall at the cost of a larger candidate list, while a higher threshold produces a shorter, more enriched list. In our validation of the classifier’s recall against a rare disease cohort (CRDC) where 2,049 patients are eligible for scoring, the OR for the PU-trained model among rule-unflagged patients remains stable between 5.5 and 6.5 across most cutoffs and rises further at calibrated probability cutoff 0.3. This consistency indicates that the PU-classifier captures a durable rare-disease signal in diagnostic history missed by the rule-based algorithm. Among rule-flagged patients, the OR starts with only 3.3 at cutoff 0, indicating a moderate enrichment by the rule-based algorithm alone, which then climbs steadily to a peak of around 6.5 at cutoff 0.4, suggesting that the addition of a high model probability substantially improves the concentration of true rare-disease patients within the rule-flagged group. Combining both approaches therefore allows RESCUE to achieve greater enrichment than either approach alone. These patterns suggest a natural prioritization ladder: when the capacity is limited, institutions may first prioritize the rule-flagged, high-probability intersection for a short, high-confidence list, while those with broader outreach infrastructure can extend to the model-only stratum where both enrichment and candidate volume is higher.

We initially started our design without providing validated Python script or Snowflake query examples and observed that the agent behavior was difficult to control and hard to reproduce^31^. This is likely because human instruction is more ambiguous than actual program codes. To ensure the reproducibility of the agent’s output, we provided agents a set of manually validated query examples as starting scripts. One limitation is that RESCUE has not been tested in across institutions. Cross-site deployment of LLM-based clinical systems faces regulatory barriers, especially different institutional policies on granting HIPAA-compliant LLMs access to patient databases that make multi-site generalization a nontrivial undertaking than a simple replication exercise. Challenges remain in regulating HIPAA-compliant LLMs and governing AI agents that connect to internal clinical data warehouses across different institutions. However, agents should be able to generate institution-specific database queries given institution-specific examples, since the goals are clearly defined in the skills we provided. The framework is also not tied to a single LLM provider and should be adapted to other agentic infrastructure. Overall, RESCUE is best viewed as a step toward automating local implementation of screening workflows, not as a replacement for validated algorithms.

## Declaration of generative AI in scientific writing

AI-assisted technologies (Claude, ChatGPT) were used during the preparation of this manuscript for proofreading and language revision. The authors reviewed and edited all AI-generated suggestions and took full responsibility for the final content of the manuscript.

## Supporting information

Prompt library, Figure S1, Figure S2, Table S1, Table S2, Table S3, Table S4

## Data Availability

All data produced are available online at https://github.com/stormliucong/RESCUE-Agent

## Acknowledgements

This study was funded by the National Institute of Health / National Human Genome Research Institute (NIH / NHGRI) under award number R01HG120566.

## Author Contributions

W.C. and C.L. conceptualized the idea; C.L. conducted the study, analysis, and drafted the manuscript; C.F. provided CRDC data; A.A. and M.N. edited the manuscript; A.G. conducted manual chart review.

## Data and code availability

The data used in this study contain patient-sensitive information and will, therefore, not be published. Requests for access to de-identified data may be directed to the corresponding author and are subject to institutional review and a data use agreement with Boston Children’s Hospital.

The source code is available at https://github.com/stormliucong/RESCUE-Agent.

## Publication Ethics and Informed Consent

This study involves real patient data from Boston Children’s Hospital and was approved by the hospital’s institutional review board.

## References

1. Nguengang Wakap S, Lambert DM, Olry A, et al. Estimating cumulative point prevalence of rare diseases: analysis of the Orphanet database. Eur J Hum Genet 2020;28:165–73. DOI: 10.1038/s41431-019-0508-0.

2. Yang Y, Muzny DM, Reid JG, et al. Clinical whole-exome sequencing for the diagnosis of mendelian disorders. N Engl J Med 2013;369:1502–11. DOI: 10.1056/NEJMoa1306555.

3. Clark MM, Stark Z, Farnaes L, et al. Meta-analysis of the diagnostic and clinical utility of genome and exome sequencing and chromosomal microarray in children with suspected genetic diseases. NPJ Genom Med 2018;3:16. DOI: 10.1038/s41525-018-0053-8.

4. 100,000 Genomes Project Pilot Investigators, Smedley D, Smith KR, et al. 100,000 Genomes Pilot on rare-disease diagnosis in health care — preliminary report. N Engl J Med 2021;385:1868–80. DOI: 10.1056/NEJMoa2035790.

5. Manickam K, McClain MR, Demmer LA, et al. Exome and genome sequencing for pediatric patients with congenital anomalies or intellectual disability: an evidence-based clinical guideline of the American College of Medical Genetics and Genomics (ACMG). Genet Med 2021;23:2029–37. DOI: 10.1038/s41436-021-01242-6.

6. Malinowski J, Miller DT, Demmer L, et al. Systematic evidence-based review: outcomes from exome and genome sequencing for pediatric patients with congenital anomalies or intellectual disability. Genet Med 2020;22:986–1004. DOI: 10.1038/s41436-020-0771-z.

7. Bauskis A, Strange C, Molster C, Fisher C. The diagnostic odyssey: insights from parents of children living with an undiagnosed condition. Orphanet J Rare Dis 2022;17:233. DOI: 10.1186/s13023-022-02358-x.

8. Splinter K, Adams DR, Bacino CA, et al. Effect of genetic diagnosis on patients with previously undiagnosed disease. N Engl J Med 2018;379:2131–39. DOI: 10.1056/NEJMoa1714458.

9. Wei WQ, Bastarache LA, Carroll RJ, et al. Evaluating Phecodes, clinical classification software, and ICD-9-CM codes for phenome-wide association studies in the electronic health record. PLoS One 2017;12:e0175508. DOI: 10.1371/journal.pone.0175508.

10. Köhler S, Gargano M, Matentzoglu N, et al. The Human Phenotype Ontology in 2021. Nucleic Acids Res 2021;49:D1207–D1217. DOI: 10.1093/nar/gkaa1043.

11. Zhao M, Havrilla JM, Fang L, et al. Phen2Gene: rapid phenotype-driven gene prioritization for rare diseases. NAR Genom Bioinform 2020;2:lqaa032. DOI: 10.1093/nargab/lqaa032.

12. Zhai W, Huang X, Shen N, Zhu S. Phen2Disease: a phenotype-driven model for disease and gene prioritization by bidirectional maximum matching semantic similarities. Brief Bioinform 2023;24:bbad172. DOI: 10.1093/bib/bbad172.

13. Johnson R, Stephens AV, Mester R, et al. Electronic health record signatures identify undiagnosed patients with common variable immunodeficiency disease. Sci Transl Med 2024;16:eade4510. DOI: 10.1126/scitranslmed.ade4510.

14. Köhler S, Schulz MH, Krawitz P, et al. Clinical diagnostics in human genetics with semantic similarity searches in ontologies. Am J Hum Genet 2009;85:457–64. DOI: 10.1016/j.ajhg.2009.09.003.

15. Yang H, Robinson PN, Wang K. Phenolyzer: phenotype-based prioritization of candidate genes for human diseases. Nat Methods 2015;12:841–3. DOI: 10.1038/nmeth.3484.

16. Smedley D, Jacobsen JOB, Jäger M, et al. Next-generation diagnostics and disease-gene discovery with the Exomiser. Nat Protoc 2015;10:2004–15. DOI: 10.1038/nprot.2015.124.

17. Birgmeier J, Haeussler M, Deisseroth CA, et al. AMELIE speeds Mendelian diagnosis by matching patient phenotype and genotype to primary literature. Sci Transl Med 2020;12:eaau9113. DOI: 10.1126/scitranslmed.aau9113.

18. Ediae GU, White-Brown A, Chisholm C, et al. ThinkRare: a search algorithm to identify patients with undiagnosed rare genetic disease in an electronic medical record. Genet Med 2025;27:101570. DOI: 10.1016/j.gim.2025.101570.

19. Singhal K, Azizi S, Tu T, et al. Large language models encode clinical knowledge. Nature 2023;620:172–80. DOI: 10.1038/s41586-023-06291-2.

20. Groza T, Caufield H, Gration D, et al. An evaluation of GPT models for phenotype concept recognition. BMC Med Inform Decis Mak 2024;24:30. DOI: 10.1186/s12911-024-02439-w.

21. Chen X, Faviez C, Vincent M, Saunier S, Garcelon N, Burgun A. Improving patient similarity using different modalities of phenotypes extracted from clinical narratives. Stud Health Technol Inform 2023;302:1037–41. DOI: 10.3233/SHTI230342.

22. Yao S, Zhao J, Yu D, et al. ReAct: synergizing reasoning and acting in language models. In: Proceedings of the 2023 International Conference on Learning Representations. Kigali, Rwanda: ICLR, 2023. (https://arxiv.org/abs/2210.03629)

23. Schick T, Dwivedi-Yu J, Dessì R, et al. Toolformer: language models can teach themselves to use tools. In: Proceedings of the 2023 Conference on Neural Information Processing Systems. New Orleans, LA: NeurIPS, 2023. (https://arxiv.org/abs/2302.04761)

24. Murphy SN, Weber G, Mendis M, et al. Serving the enterprise and beyond with informatics for integrating biology and the bedside (i2b2). J Am Med Inform Assoc 2010;17(2):124–130. DOI: 10.1136/jamia.2009.000893.

25. Rockowitz S, LeCompte N, Carmack M, et al. Children’s rare disease cohorts: an integrative research and clinical genomics initiative. NPJ Genom Med 2020;5:29. DOI: 10.1038/s41525-020-0137-0.

26. Lewis FL, Zhang H, Hengster-Movric K, Das A. Cooperative control of multi-agent systems: optimal and adaptive design approaches. London: Springer, 2014.

27. Banerjee J, Taroni JN, Allaway RJ, Prasad DV, Guinney J, Greene C. Machine learning in rare disease. Nat Methods 2023;20:803–14. DOI: 10.1038/s41592-023-01886-z.

28. Dagli MM, Ghenbot Y, Ahmad HS, et al. Development and validation of a novel AI framework using NLP with LLM integration for relevant clinical data extraction through automated chart review. Sci Rep 2024;14:26783. DOI: 10.1038/s41598-024-77535-y.

29. Wu P, Gifford A, Meng X, et al. Mapping ICD-10 and ICD-10-CM codes to Phecodes: workflow development and initial evaluation. JMIR Med Inform 2019;7:e14325. DOI: 10.2196/14325.

30. Devlin J, Chang M-W, Lee K, Toutanova K. BERT: pre-training of deep bidirectional transformers for language understanding. In: Proceedings of the 2019 Conference of the North American Chapter of the Association for Computational Linguistics: Human Language Technologies. Minneapolis, MN: Association for Computational Linguistics, 2019:4171–86. DOI: 10.18653/v1/N19-1423.

31. Xiang Y, Yan H, Ouyang S, Gui L, He Y. SciReplicate-Bench: benchmarking LLMs in agent-driven algorithmic reproduction from research papers. DOI: 10.48550/arXiv.2504.00255.

