## Supplementary material for "RESCUE: An end-to-end multi-agent LLM system for proactive rare-disease patient screening in the EHR": Prompt library, Figure S1, Figure S2, Table S1, Table S2, Table S3, Table S4

**Supplementary Appendix**

Contents

- Prompt Library: system prompt and agent prompts (P0~P8)
- Agent-prompt map
- Supplementary Tables S1–S5
- Supplementary Figure S1, S2

**Prompt Library**

All prompts below were executed against **gpt-4.1-mini** (Azure OpenAI, temperature = 0, JSON-mode where supported). Agent assignments map each prompt/context to the agent that issues it (see *Methods*).

***P0. System Prompt (for each agent)***

You are an autonomous software engineering agent maintaining the RESCUE rare-disease EHR screening pipeline. You are responsible for ONE pipeline agent and EVERY stage it owns: generate a validated, runnable script under generated/ for each stage, by reading and adapting that stage's original skill script.

AGENT {agent.code} ({agent.name})

{agent.description}

STAGES YOU OWN — generate and validate ALL of them, in this order:

{stages_doc}

WORKFLOW (use your tools; handle the stages one at a time, in order):

1. read_file the stage's original skill script; read any rescue_common/config modules it imports (use list_dir to discover sibling helper modules in its skill folder).

2. write_file the (possibly modified) module to its generated/ path above.

3. run_command `{PYTHON} -m py_compile generated/<name>.py` and fix any errors. You MAY also run it with its flags to dry-run/verify behavior, but DB access needs the VPN.

4. Work step by step with tool calls. If you get stuck, read the error messages carefully and iterate on the generated code until all stages are validated. You can ask the user for clarification or help interpreting errors if needed.

5. Always call finish_stage() after validating each stage. When all stages are done, reply with DONE.

HARD CONTRACT (never break it, for every stage):

- Keep each script's command-line interface EXACTLY (same argparse flags incl. --dry-run and --eval-variant choices) and the same entrypoint.

- Keep the files it reads and the output paths it writes; keep importing shared helpers from rescue_common and constants from config (SCREENING_DATES, EVAL_VARIANTS, resolve_eval_variant) rather than hardcoding them.

- Preserve all SQL, LLM prompts, numeric thresholds, random seeds and clinical logic UNLESS the user instruction below tells you to change them or you find a clear need to change them.

USER INSTRUCTION:

{instruction or '(none)'}

***P1. Ontology Agent: ICD code classification***

*Used by:* Ontology Agent (Methods §"Ontology Agent")

Your task is to classify an ICD code as 'Genetic Diagnosis', 'Genetic Indication', or 'Non genetics'.

- A **Genetic Diagnosis** means the diagnosis is established based on genetic testing basis. (e.g. "Chromosome inversion in normal individual", "Klinefelter syndrome", "Marfan syndrome with skeletal manifestations")
- A **Genetic Indication** means it could indicate a strong need for genetic testing for undiagnosed rare diseases; (e.g. "Congenital malformation syndromes predominantly associated with short stature", "Gastroschisis")
- **Non genetics** means it does not significantly associated with genetics (e.g. "Toxic effect of ketones", "Low-tension glaucoma")

Return ONLY a JSON object: {"label": "<one of: Genetic Diagnosis, Genetic Indication, Non genetics>"}

ICD Code Description: *<inserted at runtime>*

***P2. Ontology Agent: Test code classification***

*Used by:* Ontology Agent

Your task is to classify a test code as one of the following categories based on the description provided:

- **Diagnostic Genetic Test for Rare Disorders:** diagnose rare/Mendelian disorders.
- **Cancer Oncology Test:** tumor/somatic or hereditary cancer panels.
- **Genetic Screening / Predisposition (Non-diagnostic):** predictive, carrier, or risk screening.
- **Pharmacogenomic Test:** drug-response genetics.
- **Non-human Genetic Test:** pathogen/viral genetic analysis.
- **Other Human Genetic Test:** HLA typing, identity/relationship, reproductive diagnostic.
- **No Evidence of Genetic Test:** counseling, planning, history only.
- **Non-genetic:** not related to genetics.

Return ONLY a JSON object: {"label": "<category>"}

Test Code Description: *<inserted at runtime>*

***P3. Modeling Agent: WES/WGS rare-disease context in notes***

*Used by:* Modeling Agent (note-channel case discovery)

You are a clinical note analyst. You will be given one or more excerpts from a patient's clinical notes. Each excerpt is a context window around a keyword mention of genome/exome sequencing (WGS, WES, genome sequencing, exome sequencing). The matched keyword is wrapped in **double asterisks**.

**Your task:** Considering ALL the excerpts together, determine whether ANY mention of WGS/WES/genome sequencing/exome sequencing is in the context of *rare disease diagnosis* for this patient.

Answer **YES** if ANY excerpt indicates sequencing in the context of:

- Diagnosing or evaluating a suspected rare/genetic disease
- History workup for unexplained phenotypes suspicious for rare disease
- Identifying a causative genetic variant for the patient's condition
- Answer **NO** if ALL excerpts indicate sequencing only in the context of:
- Cancer/tumor genomic profiling or somatic mutation testing
- Pharmacogenomics
- Family/carrier screening without suspicion of rare disease in THIS patient
- General discussion or patient education not specific to THIS patient
- Respond with ONLY a JSON object: {"context": "yes"} or {"context": "no"}

***P4. Review Agent: Pre-screen prior-testing re-ranking***

*Used by:* Review Agent (Methods §"Review Agent")

You are a clinical note analyst reviewing patient records for evidence of prior genetic testing.

You will receive excerpts from a patient's clinical notes. Determine if there is evidence that this patient has already:

- Had genetic diagnostic testing performed (WES, WGS, exome sequencing, genetic panel, etc.). Do not include carrier screening or non-diagnostic genetic tests like microarray or Fragile X.
- Had genetic diagnostic testing ordered or recommended (possibly declined by patient/family); do not include carrier screening or non-diagnostic genetic tests like microarray or Fragile X. IMPORTANT: when there is no evidence of a performed test or an established diagnosis, an order or recommendation counts as prior testing ONLY IF the same test was ordered or recommended more than twice on different dates at least 30 days apart; an isolated, one-off order does NOT count and is treated as no prior testing.
- Has a known genetic diagnosis already established.

Answer with a JSON object: {"prior_testing": "yes" or "no"}

***P5. Evaluation Agent: Post-screen genetic work-up detection***

*Used by:* Post-Index Review Agent (Evaluation §"Post-Index Review Agent")

You are a clinical note analyst reviewing patient records for evidence of genetic testing AFTER a specific screening date. You will receive excerpts from a patient's clinical notes dated AFTER their screening date. Determine if there is evidence that this patient has:

- Had genetic diagnostic testing performed (WES, WGS, exome sequencing, genetic panel, etc.) after the screening date.
- Had genetic diagnostic testing ordered or recommended after the screening date.
- Has received a new genetic diagnosis after the screening date. (Carrier/predictive screening, prenatal aneuploidy, and pharmacogenomics are excluded from "yes".)

Answer with a JSON object: {"post_testing": "yes" or "no"}

***P6. Pre-screen ES/GS-indicated phenotype (medical-necessity judge)***

*Used by:* Review Agent (Stage 09, production) and Evaluation Agent (Stage 12, evaluation)

You are a clinical geneticist reviewing de-identified clinical notes for ONE patient, all dated before a given screen date. Decide whether this patient shows clinical evidence aligned with guidelines for whole-exome or whole-genome sequencing (WES/WGS), i.e., is a plausible candidate who could benefit from WES/WGS. Relevant signals include multiple congenital anomalies, unexplained developmental delay or intellectual disability, suspected Mendelian/rare disease, a family history suggestive of a genetic condition, or a phenotype workup that has not yielded a diagnosis. Exclude presentations that can be explained by common, well-documented conditions (e.g. prematurity, birth trauma, common aneuploidies, or drug exposure).

Respond with ONLY a single JSON object of the form: {"evidence": "yes" | "no", "explanation": "<1-3 sentences citing specific findings from the notes>"}

***P7. Chart-Review Agent: Pre-screen prior genetic testing***

*Used by:* Post-Index Review Agent (Stage 12, evaluation only)

You are a clinical geneticist reviewing de-identified clinical notes for ONE patient, all dated before a given screen date. Decide whether the patient had ANY genetic diagnostic test prior to this screen date. Genetic diagnostic tests include karyotype, FISH, chromosomal microarray (CMA/aCGH), single-gene sequencing, gene panels, exome sequencing (WES), genome sequencing (WGS), targeted variant testing, or mitochondrial testing ordered for diagnostic purposes. Do NOT count carrier screening, prenatal aneuploidy screening, or pharmacogenomics.

Respond with ONLY a single JSON object of the form: {"genetic_dx": "yes" | "no", "reference": "<short quote or paraphrase from the notes>"}

***P8. Chart-Review Agent: Post-screen genetic work-up confirmation***

*Used by:* Post-Index Review Agent (Stage 12, evaluation only)

You are a clinical geneticist reviewing de-identified clinical notes for ONE patient, all dated after a given screen date. Decide whether the patient underwent any genetic diagnostic workup after that date. A workup means an ordered genetic diagnostic test (karyotype, CMA, gene panel, WES, WGS, single-gene sequencing, targeted variant, etc.), a genetics clinic referral or visit, or documented genetic counseling for diagnostic purposes. Do NOT count carrier screening, prenatal aneuploidy screening, or pharmacogenomics.

Respond with ONLY a single JSON object of the form: {"genetic_workup": "yes" | "no", "reference": "<short quote or paraphrase from the notes>"}

**Agent → prompt map**

| **Agent** | **Prompts** |
| --- | --- |
| Ontology Agent | P1, P2 |
| Modeling Agent (PU-Learning) | *P3 (ES/GS note judge)* |
| Screening Agent | *(no LLM prompt; tool use only — Python code generation for model inference + rule-based algorithm)* |
| Review Agent | P4, P6 |
| Evaluation Agent | P5 (and P6~P8 for the chart-review sub-task, evaluation only) |

**Supplementary Tables**

**Table S1. Number of unique codes assigned to each tier by the Ontology Agent**

| **Tier** | **N unique codes** | **Tier Description** | **Role** | **Examples** |
| --- | --- | --- | --- | --- |
| Tier 1A | 1,094 | ICD codes for genetic indications | Risk Enrichment | G91.0: “Communicating hydrocephalus” |
| Tier 1B | 143 | Internal lab/proc/note codes for non-diagnostic / screening / oncology | Control Purification | LAB:3726031: "Chromosome Analysis, Solid Tumor" |
| Tier 2A | 250 | Internal lab/proc/note codes for diagnostic genetic procedure | Case defining | LAB:645059009: “Whole Exome Sequencing (Proband)” |
| Tier 2B | 452 | ICD codes for genetic diagnosis | Case defining | G12.0: “Infantile spinal muscular atrophy, type I [Werdnig-Hoffman]“ |

**Table S2. Odds ratios of the Phecode-based PU model flagged individuals at calibrated cutoffs**

| **Cutoff** | **Cases flagged** | **Risk-enriched flagged** | **Purified controls flagged** | **OR case vs. purified** | **OR risk vs. purified** |
| --- | --- | --- | --- | --- | --- |
| 0.05 | 1,109 (90.2%) | 280 (92.7%) | 6,258 (56.6%) | 7.01 | 9.56 |
| 0.10 | 799 (65.0%) | 183 (60.6%) | 2,218 (20.1%) | 7.38 | 6.12 |
| 0.15 | 675 (54.9%) | 133 (44.0%) | 1,329 (12.0%) | 8.90 | 5.76 |
| 0.20 | 585 (47.6%) | 100 (33.1%) | 915 (8.3%) | 10.05 | 5.50 |
| 0.25 | 434 (35.3%) | 63 (20.9%) | 440 (4.0%) | 13.15 | 6.39 |
| 0.30 | 427 (34.7%) | 60 (19.9%) | 422 (3.8%) | 13.40 | 6.28 |
| 0.40 | 288 (23.4%) | 32 (10.6%) | 156 (1.4%) | 21.33 | 8.37 |
| 0.50 | 159 (12.9%) | 10 (3.3%) | 44 (0.4%) | 36.85 | 8.89 |
| 0.60 | 153 (12.4%) | 10 (3.3%) | 39 (0.4%) | 39.75 | 10.02 |
| 0.70 | 64 (5.2%) | 3 (1.0%) | 6 (0.05%) | **94.0** | 19.87 |
| 0.80 | 46 (3.7%) | 2 (0.7%) | 1 (0.01%) | **289.4** | 61.33 |
| 0.90 | 35 (2.85%) | 1 (0.3%) | 0 (0%) | **656.8** | 110.0 |

**Table S3. Overlap between calibrated-probability cutoff breakdown and rule-based flags**

| **Cutoff** | **N model-positive** | **N model+ ∩ Rule-flagged** | **% Rule-flagged overlap** |
| --- | --- | --- | --- |
| 0.05 | 93,869 | 2,630 | 2.8% |
| 0.10 | 32,609 | 2,053 | 6.3% |
| 0.15 | 20,523 | 1,620 | 7.9% |
| 0.20 | 15,007 | 1,361 | 9.1% |
| 0.25 | 7,812 | 871 | 11.1% |
| 0.30 | 7,582 | 854 | 11.3% |
| 0.40 | 3,558 | 423 | 11.9% |
| 0.50 | 1,508 | 152 | 10.1% |
| 0.60 | 1,353 | 132 | 9.8% |
| 0.70 | 460 | 31 | 6.7% |
| 0.80 | 248 | 6 | 2.4% |
| 0.90 | 181 | 3 | 1.7% |

**Table S4. Recall and Odds Ratio evaluated using an independent eligible CRDC sequenced cohort based on different cut-offs**

| **Rule-flagged** | **Cutoff (>=prob)** | **N flagged**  **(total: 175,842)** | **N CRDC**  **(total: 2,049)** | **Recall** | **OR** |
| --- | --- | --- | --- | --- | --- |
| FALSE | 0 | 175,842 | 2,049 | 1 | n/a |
| FALSE | 0.05 | 93,869 | 1,795 | 0.876 | 6.272 |
| FALSE | 0.1 | 32,609 | 1,151 | 0.5617 | 5.799 |
| FALSE | 0.15 | 20,523 | 877 | 0.428 | 5.871 |
| FALSE | 0.2 | 15,007 | 709 | 0.346 | 5.902 |
| FALSE | 0.25 | 7,812 | 429 | 0.2094 | 5.969 |
| FALSE | 0.3 | 7,582 | 420 | 0.205 | 5.999 |
| FALSE | 0.4 | 3,558 | 240 | 0.1171 | 6.816 |
| FALSE | 0.5 | 1,508 | 119 | 0.0581 | 7.653 |
| FALSE | 0.6 | 1,353 | 111 | 0.0542 | 7.957 |
| FALSE | 0.7 | 460 | 44 | 0.0215 | 9.146 |
| FALSE | 0.8 | 248 | 24 | 0.0117 | 9.184 |
| FALSE | 0.9 | 181 | 12 | 0.0059 | 6.052 |
| TRUE | 0 | 2,678 | 96 | 0.0469 | 3.259 |
| TRUE | 0.05 | 2,630 | 96 | 0.0469 | 3.322 |
| TRUE | 0.1 | 2,053 | 84 | 0.041 | 3.73 |
| TRUE | 0.15 | 1,620 | 74 | 0.0361 | 4.175 |
| TRUE | 0.2 | 1,361 | 71 | 0.0347 | 4.8 |
| TRUE | 0.25 | 871 | 48 | 0.0234 | 5.042 |
| TRUE | 0.3 | 854 | 48 | 0.0234 | 5.148 |
| TRUE | 0.4 | 423 | 29 | 0.0142 | 6.318 |
| TRUE | 0.5 | 152 | 9 | 0.0044 | 5.357 |
| TRUE | 0.6 | 132 | 9 | 0.0044 | 6.229 |
| TRUE | 0.7 | 31 | 2 | 0.001 | 5.854 |
| TRUE | 0.8 | 6 | 1 | 0.0005 | 16.971 |
| TRUE | 0.9 | 3 | 1 | 0.0005 | 42.429 |

**Table S5. Evaluation based on LLM and structured codes using post-screen EHR data in different arms**

| **Arm** | **N candidates** | **N with adequate follow-up** | **Structured evidence (Tier 2A/2B)** | **Unstructured (LLM P5)** | **Union (any work-up)** | **% of follow-up** |
| --- | --- | --- | --- | --- | --- | --- |
| M+T+ | 121 | 117 (96.7%) | 29 (24.8%) | 7 (6.0%) | 31 | 26.5% |
| M-T+ | 528 | 468 (88.6%) | 27 (5.8%) | 10 (2.1%) | 33 | 7.1% |
| M+T- | 796 | 698 (87.7%) | 120 (17.2%) | 20 (4.3%) | 132 | 18.9% |
| M-T- | 968 | 671 (69.3%) | 20 (3.0%) | 7 (1.0%) | 24 | 3.6% |

**Supplementary Figures**

**Figure S1. ROC and Precision-Recall curves**

Receiver Operating Curve and precision-recall curves on the 12,591-patient holdout for the phecode and embedding PU-XGBoost classifiers.

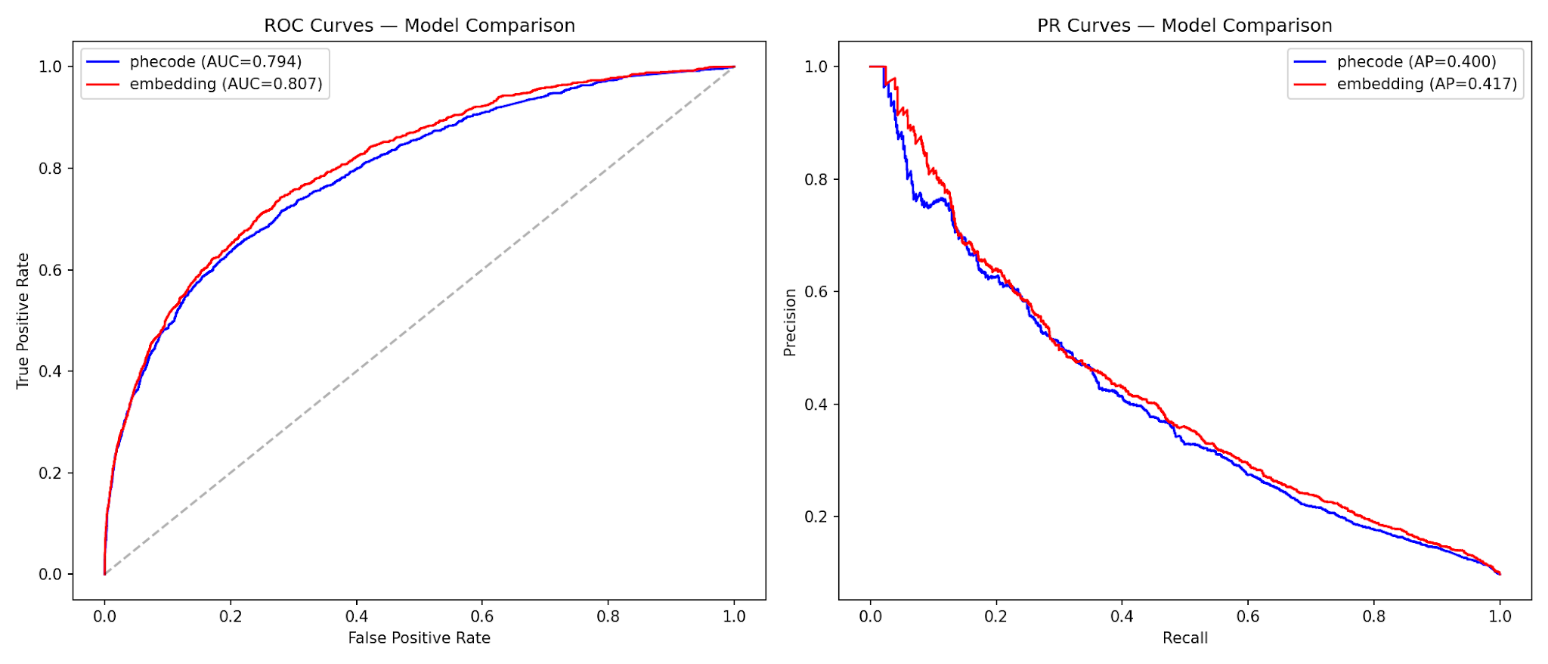
**Figure S2. Calibration plot**

Reliability diagram (predicted vs. observed positive fraction) for the isotonic-calibrated phecode and embedding models on the holdout.

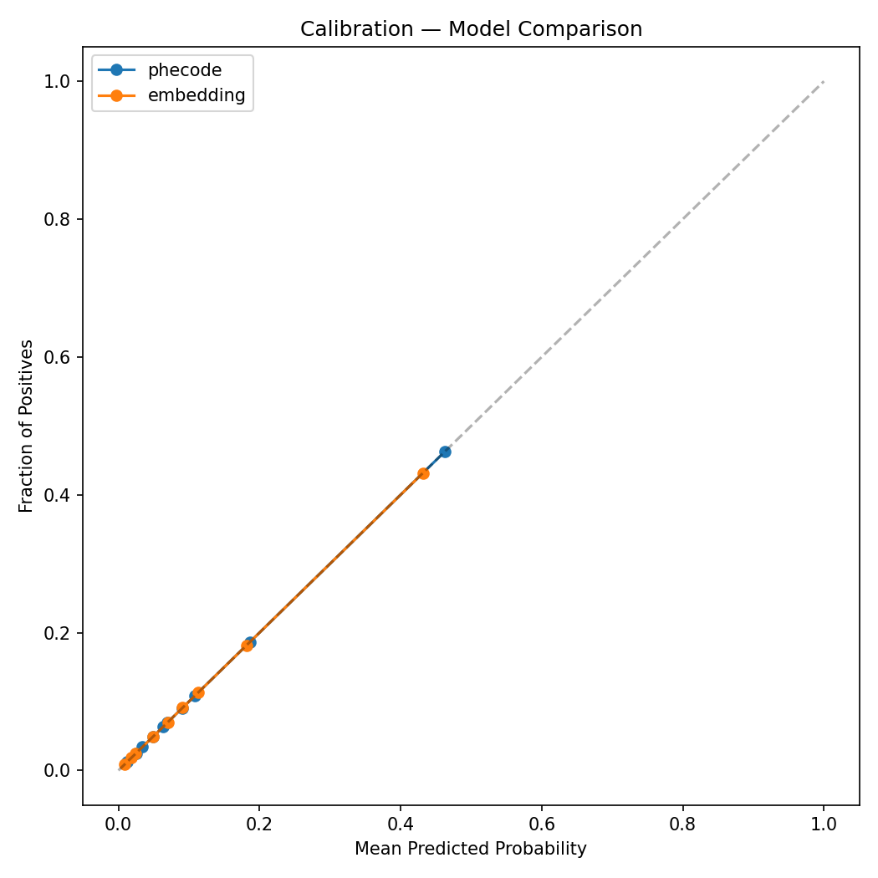
